# Pre- and Post-Flavor Ban Oral Nicotine Pouches: A Chemical, Sensory, and Young Adult Appeal Analysis

**DOI:** 10.64898/2025.12.10.25341992

**Authors:** Natalia Peraza, Ravindra Singh Thakur, Sairam V. Jabba, Jacob Aguilera, Paul W. Martines, Marielle C. Brinkman, Sven-Eric Jordt, Ahmad El Hellani, John R. Monterosso, Alayna P. Tackett

## Abstract

**Importance:** The tobacco industry exploits legislative loopholes by introducing products with novel constituents, sensory features, and/or branding to bypass flavor restrictions and maintain appeal, particularly among young populations. Post California’s 2022 flavor ban, leading oral nicotine pouch (ONP) brand (ZYN) started marketing two new “unflavored” products, Classic and Original, but it is unknown if they differed from pre-ban products marketed as “flavor-ban approved” (Chill and Smooth) in their chemical composition, sensory characteristics and human appeal.

**Methods:** Nicotine, menthol, and synthetic coolants, WS-3 and WS-23, were quantified using gas chromatography-mass spectrometry. Menthol receptors (TRPM8 and TRPA1) activity was measured by calcium-microfluorimetry to assess cooling-and irritation-potential. ONP sensory attributes and appeal were assessed in young-adults (21–25Yr old) after 5-minute standardized use in a double-blind, randomized remote trial.

**Results:** “Chill” and “Classic” ONPs contained WS-3 exclusively (0.24±0.02 mg/pouch), with consistent levels across nicotine strengths; “Smooth” and “Original” contained neither menthol nor WS-3. Extracts of “Chill” and “Classic” robustly activated the cold/menthol TRPM8-receptor with similar potency and efficacy, while weakly activating the sensory irritant TRPA1 receptor. Human participants rated “Chill” and “Classic” as having stronger cooling sensations (p<0.05), while “Classic” was rated as more minty flavored when compared to unflavored “Smooth” and “Original” (p<0.005) ONPs.

**Conclusions and Relevance:** Pre- and post-ban ONPs showed similar chemical, sensory, and appeal profiles based on WS-3 presence. By using concept names and sensory cues to disguise flavors, industry exploits regulatory loopholes to sustain marketing of banned products.

**What is already known on this topic:** Oral nicotine pouches (ONPs) have gained popularity among young adults, and manufacturers have introduced products and marketed as “ban-compliant” with concept names and sensory additives that may mimic flavors despite flavor bans.

**What this study adds:** Chemical, receptor, and sensory analyses of pre- and post-ban ZYN ONPs showed both contained the synthetic coolant WS-3, producing flavor-like cooling sensations even without menthol or other flavor compounds.

**How this study might affect research, practice, or policy:** Findings indicate that post-ban ONPs retain the same sensory and chemical properties as pre-ban products, highlighting enforcement challenges when odorless sensory additives and branding strategies allow products to retain flavor-like effects despite legislative restrictions.

## INTRODUCTION

With more than a billion units sold per month in 2024,^1^ oral nicotine pouch (ONP) products have rapidly gained popularity, prevalence, and tobacco-marketplace share in the United States of America (USA).^2^ Sales in the USA have increased from 163,000 units upon market introduction in 2016 to 9.5 billion units in 2023,^2,3^ with higher prevalence seen in younger and middle-aged men.^4,5^ In high-school adolescents, a continuous increase in ONP use is seen over the past years, increasing from 1.1% to 2.4% from 2021-2024.^6^ Analyzing a 2021 cohort of Southern California high-school adolescents and 2024 NYTS data,^7^ ONPs were demonstrated to be the 2^nd^ most prevalent nicotine product, after e-cigarettes.^8^ Among young adults (YAs) aged 21-24 in the USA, ONPs have higher current use rates^5,9,10^ and evidence indicates that awareness and favorable perceptions of ONPs are high among young adults.^11-13^ ONPs use among youth was also associated with use of other tobacco products, with current use more prevalent among Hispanics, Non-Hispanic Blacks, and those with lower education and annual incomes.^5^

ONPs are available in several youth-appealing flavors,^10,14^ with fruit (berry, citrus) and mint-/menthol-(peppermint, spearmint, wintergreen) being the most commonly used and preferred flavors.^3,15^ More recently, synthetic cooling agents containing ice-flavored ONPs have been introduced, where coolants like Wilkinson Sword-3 (WS-3) are added either to provide more intense cooling notes to flavored ONPs or to circumvent “characterizing flavor” definitions included in the local and statewide restrictions and regulations on flavored tobacco and nicotine products.^16-18^ These synthetic cooling agents, similar to menthol, elicit robust cooling sensations through activation of the cold/menthol receptor TRPM8 expressed in the trigeminal neurons innervating the buccal and oral cavities.^16,19^ However, unlike menthol, these cooling agents lack the characteristic minty odor and are much less irritating than menthol due to their reduced activity at the sensory-irritant receptor TRPA1.^16,19^ This combination of properties of eliciting cooling sensations with little to no sensory irritation has made these synthetic cooling agents a popular substitution of menthol in a wide variety of tobacco products, including e-cigarettes, ONPs and even combustible cigarettes.^16,20,21^

California’s 2022 flavor ban prohibited the sale of flavored tobacco products (with exceptions to hookah, premium cigars, and looseleaf pipe tobacco)^22^ to reduce youth appeal, initiation, and tobacco-related health disparities.^22^ The legislation defined “characterizing flavors” as any natural or artificial taste or aroma beyond tobacco and nicotine.^22^ Still, some tobacco companies appear to circumvent flavor restrictions by marketing products with ambiguous names (e.g., “clear,” “non-menthol”) and color-coded packaging to suggest cooling sensations without overtly signaling flavor.^20,23-25^ These tactics highlight challenges enforcing a comprehensive flavor ban and underscore the need for research on how modifications in chemical composition, sensory attributes, and marketing affect product appeal.

ZYN, the leading U.S. oral nicotine pouch (ONP) brand^26^–a product class with increasing popularity among young people^27^–strategically marketed their products in California prior to implementation of the 2022 flavor ban using non-characterizing flavor messaging. ZYN advertised ONPs “Chill” and “Smooth” as “flavor-ban approved”, yet “Chill” was demonstrated to contain a synthetic coolant, WS-3, and consumer reviews suggested it still provided a cooling sensation, raising questions about compliance.^18^ ZYN withdrew the “Chill” and “Smooth” varieties in California after the ban, and released “Classic” and “Original” ONPs instead, avoiding explicit flavor references (e.g., Chill). However, “Classic” employs blue packaging similar to “Chill” that might still signal a cooling sensation to consumers. It is unknown whether the post-ban ONPs align with or deviate from the chemical makeup, sensory attributes, and appeal of the pre-ban ONPs.

This multidisciplinary study examined the chemical characteristics, menthol receptors activity (*in vitro*), and appeal/sensory ratings among YA’s who currently use nicotine for four ZYN ONPs: two post-ban products sold in California (Original and Classic) and two pre-ban products (Chill and Smooth) purchased in non-flavor ban states.

## METHODS

### ONP Product Purchase

ZYN “Original” and “Classic” ONPs (3mg and 6mg) were obtained in Los Angeles, California, in September-November 2024, while ZYN “Smooth” and “Chill” ONPs (3mg and 6mg) were purchased in Columbus, Ohio, (December 2024) and Durham, North Carolina, (2023) from local convenience stores.

### Chemical Characterization of ONPs

Individual pouches of each ONP (n=3) were weighed and extracted in 10 mL of methanol, subjected to 30 minutes of vertical shaking followed by 30 minutes of sonication. To remove water content, 100 mg of anhydrous sodium sulfate was added, vortexed for 1 minute, and centrifuged for 5 minutes. The supernatant was then collected, filtered (0.22 µm syringe filter), diluted, and directly injected (1 μL) into a gas chromatography - mass spectrometry system (GC-MS). ONP extracts were analyzed for nicotine (CAS No. 54-11-5), menthol (89-78-1), WS-23 (51115-67-4), and WS-3 (39711-79-0) using standards from Sigma-Aldrich (purity ≥ 99%), against an internal standard quinoline (91-22-5) 1 µg/mL purchased from Thermo Fisher Scientific Chemicals (purity=98.8%).

GC-MS analysis was performed using an Agilent 8890 gas chromatograph (GC) coupled with a 5977B mass spectrometer (MS). The inlet temperature was 250°C, the MS transfer line was 260°C, and the source temperature was 230°C. Separation was achieved on a 30 m DB-BAC1 UI column (0.32 mm i.d., 1.8 μm film thickness). The temperature program started at 100°C (1 min hold), ramped at 20°C/min to 180°C, then 25°C/min to 250°C, with a 3-minute final hold. Helium, at a flow rate of 1 mL/min, was used as the carrier gas. The mass spectrometer operated in electron impact (EI) mode at 70 eV. Total run time: 10.8 minutes.^16^

### Testing for Sensory Cooling and Irritant Activity by Calcium-Microfluorimetry

#### Extract Preparation for Calcium Microfluorimetry

ZYN pre-ban (Smooth and Chill) and post-ban (Original and Classic) ONPs (3mg) and ZYN “Cool-Mint” (3mg, positive control) were extracted with calcium assay buffer (Hank’s Balanced Salt Solution with 10 mM HEPES; HBSS). Extracts were prepared by stirring pouch contents overnight in 10 mL of calcium assay buffer HBSS using a shaker. Contents were centrifuged (@4000 rpm for 5 minutes) and supernatants were collected. Further dilutions of these extracts (diluted 1X-100X in assay buffer) were prepared to test for receptor activity. 1X dilution is defined as the extract of one pouch contents in 10 mL assay buffer, and 100X is 100-fold dilution thereof.

#### Calcium-Microfluorimetry to Measure Sensory Receptor Responses

Pouch extracts (diluted 1X-100X in buffer) were tested for cold/menthol receptor, TRPM8, and sensory irritant receptor, TRPA1 activity by intracellular calcium microfluorimetry in HEK-293t cells (RRID:CVCL1926) expressing human TRPM8 and TRPA1 isoforms and as previously described.^16,20^ To control for any variability in receptor expression levels and loading of Ca^2+^ indicator across experiments, Ca^2+^-influx responses from these extracts were normalized to the Ca^2+^-response elicited by a maximally activating concentration of specific agonists L-menthol (1 mM; TRPM8) and cinnamaldehyde (CAD; 1 mM; TRPA1). Corresponding nicotine concentrations were utilized as background controls.

### Sensory and Appeal Procedures

#### Participants

Eligibility criteria were: (1) aged 21-25, (2) past 30-day e-cigarette use and/or cigarette use,^14^ (3) access to a location where study sessions can be conducted without interruption (e.g., residence), (4) access to an electronic device (e.g., laptop, tablet, smart phone) and consistent internet connection to conduct online study sessions via Zoom, and (5) willingness to try ONPs. Exclusion criteria were: (1) having a plan to reduce or quit vaping or smoking in the next month, (2) current use of smoking cessation medication (e.g., Nicotine Replacement Therapy), (3) use of an ONP more than 5 times in the last 30 days (to avoid effects of recent exposure to the ONP study product class), (4) daily use of other nicotine products (i.e., other than e-cigarettes or cigarettes), (5) being pregnant or breastfeeding, and (6) medical conditions (e.g., cardiovascular or lung disease) that are contraindications to nicotine use.

#### Design and Procedure

Following eligibility confirmation and successful shipping of study products, participants attended a 2-hour virtual video-conferenced laboratory session applying procedures successfully used for virtual studies of e-cigarette and oral nicotine product appeal previously.^14,28,29^ The double-blind test procedure during the remote laboratory session involved self-administration of 4 different ZYN ONPs; all products included 3mg nicotine strength. The 4 ZYN ONPs (Classic, Original, Chill, Smooth) were presented in a randomized order, constituting a within-subjects design where participants evaluated all product flavors of the same product type and brand. Randomization was carried out in advance by a graduate research assistant using R to generate a unique random sequence of the four products for each participant. Each sequence was generated using a reproducible random seed, and no sequence was reused across participants. During the session, experimenters implemented the assigned sequence by instructing participants to display the number printed on each product’s non-descript packaging prior to administration, ensuring adherence to the correct order. The graduate research assistant did not administer any sessions, maintaining blinding for both participants and experimenters to the product identities (i.e., double-blind). Participants completed demographic and tobacco product use surveys between product administration blocks (2 products per block, 2 blocks separated by a 20-minute period). During each administration, participants were provided with instructions by the research staff on how to use each randomized ONP. Participants were instructed: “To use this product, place it between your upper lip and gum. It fits comfortably in the mouth and does not require spitting. You will be trying this product for 5 minutes.” After instructions, participants were asked to keep the respective ONP in their mouth for 5 minutes. Each sequence was separated by a 5-minute period during which participants were asked to rinse their mouths with water to cleanse their palates and prevent sensory carryover across trials.

#### Participant Characteristics

Participants reported demographic, vaping, and smoking characteristics. The Hooked on Nicotine Checklist (HONC)^30^ for e-cigarette and/or cigarettes were also administered (range: 0-10), as appropriate (i.e., dual users got both versions of the HONC; See Supplemental Table 1 for details).

#### Outcome Measures

After each ONP administration, participants rated three dimensions of appeal and seven sensory attributes. The following items used in previous research on ONP sensory attributes and appeal were utilized.^28^ The appeal items include: (1) “How much did you like the product?” (100 mm Visual Analogue Scale [VAS], 0-100 with “Not at all” to “Extremely” as anchors) (2) “How much did you dislike the product?” (VAS, “Not at all” to “Extremely”); (3) “Would you use the product again?” (VAS, “Not at all” to “Definitely”). The sensory experience items include: (1) “How sweet was the product?” (VAS, “Not at all” to “Extremely”); (2) “How bitter was the product?” (VAS, “Not at all” to “Extremely”); (3) “How smooth was the product?” (VAS, “Not at all” to “Extremely”); (4) “How harsh was the product?” (VAS, “Not at all” to “Extremely”); (5) “How strong was the flavor of the product?” (VAS, “Not at all” to “Extremely”); (6) “How much did the product tingle in your mouth?” (VAS, “Not at all” to “Extremely”); (7) “How strong was the burning sensation in your mouth?” (VAS, “Not at all” to “Extremely”). Following a method previously used in assessing the appeal of e-cigarettes and oral nicotine products,^14,28,29^ composite appeal was calculated by averaging “Liking”, “Disliking” (reverse-coded), and “Willingness to Use Again”. Each of the remaining items were analyzed separately rather than combined into scales, allowing for a comprehensive examination of each aspect of the sensory experience across the various oral nicotine products tested in the study.

#### Data Analyses

Two-tailed unpaired t-tests were performed to assess differences in the chemical composition. Dose-response curves for receptor activity and associated calcium influx changes were plotted using non-linear regression analysis with a 4-parameter logistic equation (Graphpad Prism 10.0, San Diego, CA). Human laboratory data were analyzed in R using linear regression to examine differences in appeal and sensory ratings across four ONPs with “Smooth” as the reference. Regression coefficients, standard errors, and p-values (<0.05) determined product differences and Welch two-sample t-test compared appeal ratings between pre- and post-ban ONPs. Given the exploratory nature of the analyses and the limited number of comparisons, corrections for multiple comparisons were not applied.

## RESULTS

### Chemical Composition (Table 1)

Synthetic coolant WS-3 was detected in post-ban ONP “Classic” at similar levels to pre-ban ONP “Chill” (∼0.25mg/pouch; p>0.42), with levels being consistent between 3mg and 6mg ONPs. WS-3 was not detected in “Original and Smooth”. Also, WS-23 and menthol were not detected in any analyzed ONP. Nicotine amounts across pre- and post-ban ZYN ONPs were similar (p>0.16), except for those between “Smooth” 6mg and “Original” 6mg (5.54±0.04 vs. 6.15±0.19; p<0.005).

### Sensory Receptor Activation Assays

Extracts of WS-3 containing ONPs (Chill, Classic), and menthol containing “Cool Mint” ONP robustly activated cold/menthol TRPM8 receptors, with “Chill” and “Classic” displaying higher efficacies than “Cool Mint” at low dilutions (2X and 3.3X; Figure 1A). Responses of TRPM8-expressing cells to ZYN “Original” and “Smooth” extracts were similar to background indicating the absence of menthol or synthetic coolants. “Cool Mint” extracts strongly activated the irritant receptor TRPA1 even at 20x dilution, while “Chill” and “Classic” extracts produced weaker activity, similar to “Original” and “Smooth” ONPs (3.3X and above; Figure 1B).

**Figure 1.**
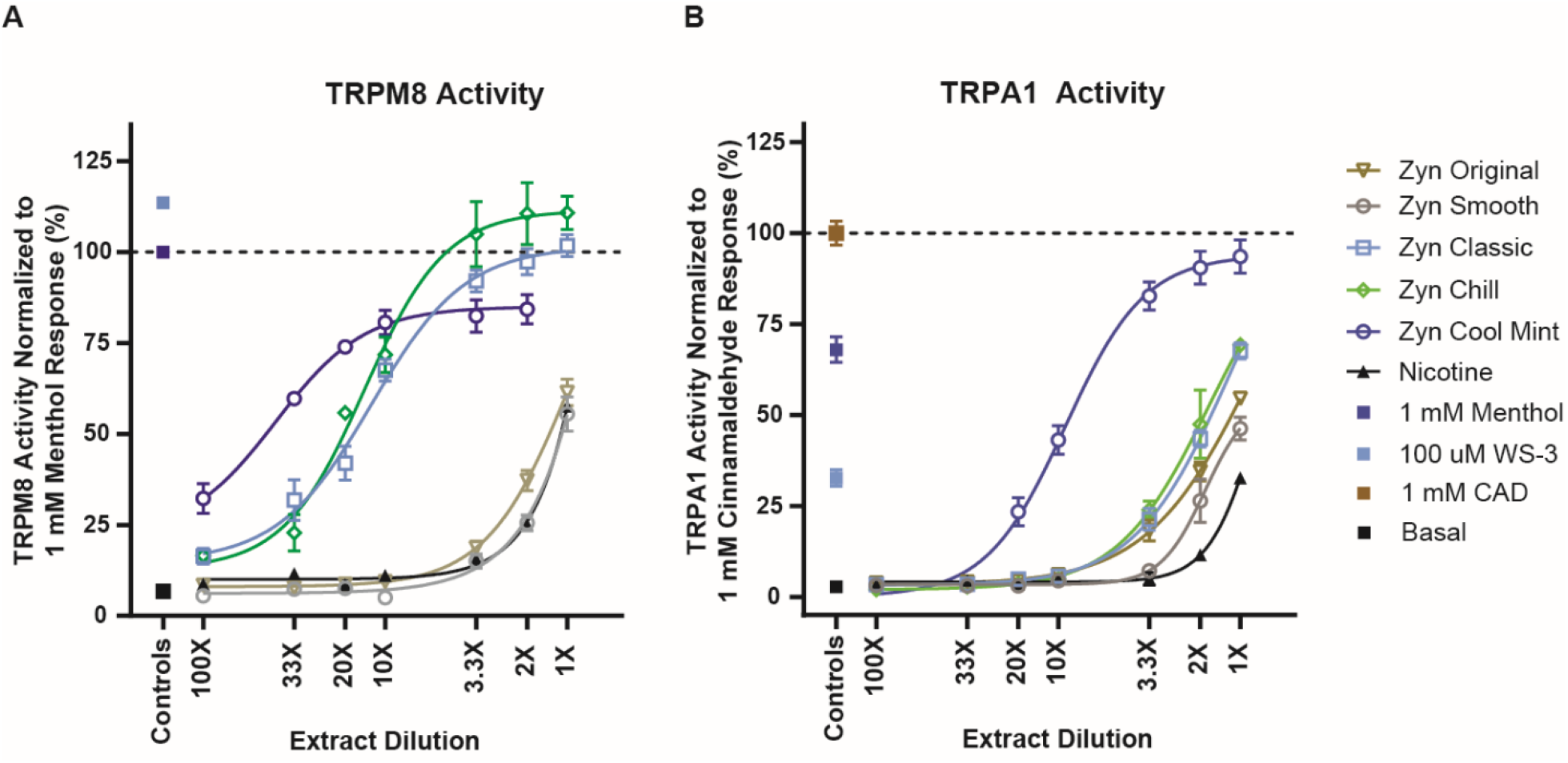
Sensory cooling and irritant activity of ZYN ONPs pre- and post-flavor ban in California *Note*. A, Dose-response analysis of human TRPM8 cold/menthol receptor-mediated calcium ion influx upon exposure to series of extract dilutions prepared from various ZYN brand ONPs. The activity of the receptor and subsequent influx of calcium ions into the cells was measured as the change in fluorescence units (F_max_□−□F_0_), was normalized to the calcium influx response elicited by a saturating concentration of TRPM8 specific agonist L-menthol (1 mM; solid blue square; horizontal black dotted line). Response to a saturating concentration of synthetic cooling agent WS-3 (100 μM; solid teal square), nicotine dose-response (solid black triangle) and vehicle control (basal; solid black square) shown for comparison. B, Dose-response analysis of human TRPA1, a sensory irritant receptor, performed as in A for ZYN brand ONPs, except that responses were normalized to the calcium influx response elicited by a saturating concentration of TRPA1 specific agonist cinnamaldehyde (CAD; 1 mM; solid orange square; horizontal black dotted line). Error bars for each data point show SE of the mean. TRPM8 indicates transient receptor potential cation channel subfamily M member 8; TRPA1 indicates Transient receptor potential cation channel subfamily A member 1.

### Human Laboratory Data

Participants (N=22; M_age_=22.8; 31.1% female) primarily reported current e-cigarette use (77.3%), with a minority reporting cigarette use or both e-cigarette and cigarette use (9.1%), and exhibited moderate levels of nicotine dependence (Table 2). “Chill” and “Classic” products were rated higher for appeal compared to “Smooth”, but these differences were not statistically significant (p>0.05; Figure 2A). “Classic” was rated significantly more minty than “Smooth” (p<0.05; Figure 2B), but no differences were observed for “Chill” (p=0.07) and “Original” (p=0.85). Both “Chill” and “Classic”, but not “Original”, were rated significantly higher than “Smooth” for cooling sensation (p<0.001 and p<0.01, respectively; Figure 2C). “Original” did not differ from “Smooth” for appeal (p=0.81) or across most sensory attributes, except for tingling sensation (p<0.05; Figure 2D). There were no significant differences in appeal between “Chill” and “Classic” (p=0.98) or “Smooth” and “Original” (p=0.81; see Table 3).

**Table 1.** Level of WS-3 and nicotine (mean ± standard deviation; n=3 replicates per brand) in ZYN ONPs pre- and post-flavor ban in California.

|  | Oral Nicotine Pouches | Concentration in mg/pouch |  |
| --- | --- | --- | --- |
|  |  | Nicotine | WS-3 |
| Pre-ban Products | ZYN Chill 3mg | 2.89 ± 0.14 | 0.25 ± 0.00 |
| | ZYN Chill 6mg | 5.61 ± 0.33 | 0.24 ± 0.03 |
| | ZYN Smooth 3mg | 2.84 ± 0.12 | ND |
| | ZYN Smooth 6mg | 5.54 ± 0.04 | ND |
| Post-ban Products | ZYN Classic 3mg | 2.88 ± 0.03 | 0.24 ± 0.02 |
| | ZYN Classic 6mg | 5.94 ± 0.07 | 0.25 ± 0.01 |
| | ZYN Original 3mg | 2.77 ± 0.11 | ND |
| | ZYN Original 6mg | 6.15 ± 0.19 | ND |

*Note.* n: represents number of replicates. ND: Not detected.

**Table 2.** Participant Characteristics (N=22)

| Variables | N (%) or M (SD) |
| --- | --- |
| Age, M (SD) | 22.8 (1.2) |
| Gender identity, N (%) |  |
| Man | 15 (68.2) |
| Woman | 7 (31.8) |
| Sexual identity, N (%) |  |
| Straight/heterosexual | 13 (59.1) |
| Another sexual identity<sup>a</sup> | 9 (40.9) |
| Race/ethnicity, N (%) |  |
| Asian | 1 (4.6) |
| Black or African American | 1 (4.6) |
| Hispanic, Latino, or Spanish origin | 4 (18.1) |
| Non-Hispanic, White | 9 (40.9) |
| Multiracial | 7 (31.8) |
| Educational attainment, N (%) |  |
| High school diploma/GED or less | 4 (18.1) |
| Some college completed or currently enrolled | 6 (27.3) |
| College degree or higher | 10 (45.5) |
| Not currently enrolled in school | 2 (9.1) |
| Employment status, N (%) |  |
| Full-time | 14 (63.6) |
| Part-time | 2 (9.1) |
| Unemployed | 1 (4.6) |
| Student | 5 (22.7) |
| Current nicotine use status N (%) |  |
| Current e-cigarette use (with no history of cigarette use) | 17 (77.3) |
| Current cigarette use (with history of e-cigarette use) | 3 (13.6) |
| Current dual use of e-cigarette and cigarette | 2 (9.1) |
| HONC: E-cigarette<sup>b</sup>, M (SD) | 7.4 (2.5) |
| HONC: Cigarette<sup>c,d</sup>, M (SD) | 5.7 (5.1) |
| No. days used e-cigarettes in past 30 days, M (SD) | 26.0 (8.4) |
| No. days used cigarettes in past 30 days, M (SD)<sup>d</sup> | 7.0 (7.1) |

*Note.* Frequencies may not sum to the total due to different patterns of missing data across variables.
<sup>a</sup> Bisexual (n=4), gay (n=3), lesbian (n=1), or questioning/unsure (n=1).
<sup>b</sup> Hooked on Nicotine Checklist: E-cigarette (range: 0-10).
<sup>c</sup> Hooked on Nicotine Checklist: Cigarette (range: 0-10).
<sup>d</sup> Among subsample that smoked cigarettes.

**Table 3.** Differences in appeal and sensory ratings across ZYN ONPs pre- and post-flavor ban in California: Regression and Pairwise Comparisons.

| Outcome Variable | Product | Estimates (b) | SE | F | t-value | p-value |
| --- | --- | --- | --- | --- | --- | --- |
| Appeal | Chill | 6.24 | 7.36 | 0.3858 | 0.85 | 0.39 |
|  | Classic | 6.47 |  |  | 0.88 | 0.38 |
|  | Original | 1.81 |  |  | 0.81 | 0.81 |
| Cooling Sensation | Chill | 23.05 | 6.66 | 6.031 | 3.46 | <0.001 *** |
|  | Classic | 19.96 |  |  | 2.99 | <0.01 ** |
|  | Original | 3.46 |  |  | 0.52 | 0.61 |
| Minty Flavor | Chill | 14.14 | 7.65 | 2.935 | 1.85 | 0.07 |
|  | Classic | 18.68 |  |  | 2.44 | 0.02 * |
|  | Original | 1.46 |  |  | 0.19 | 0.85 |
| Tingling Sensation | Chill | 18.68 | 7.69 | 2.681 | 2.43 | 0.02 * |
|  | Classic | 13.77 |  |  | 1.79 | 0.08 |
|  | Original | 19.00 |  |  | 2.47 | 0.02 * |
| Outcome Variable | Product Comparison | Estimates (B) | - | - | t-value | p-value |
| Appeal | Chill vs. Classic | -0.23 | - | - | -0.03 | 0.98 |
|  | Smooth vs. Original | 1.82 | - | - | 0.24 | 0.81 |

*Note.* Top Section of Table: Results from linear regression models with “Smooth” as the reference category. Asterisks indicate significant differences from “Smooth”. Significant effect of product on appeal or sensory ratings: \* $p < 0.05$ , \*\* $p < 0.01$ , \*\*\* $p < 0.001$ . Bottom Section of Table: Results from independent t-tests comparing pre-flavor ban products (Chill, Smooth) to their post-flavor ban product counterparts (Classic, Original).

**Figure 2.**
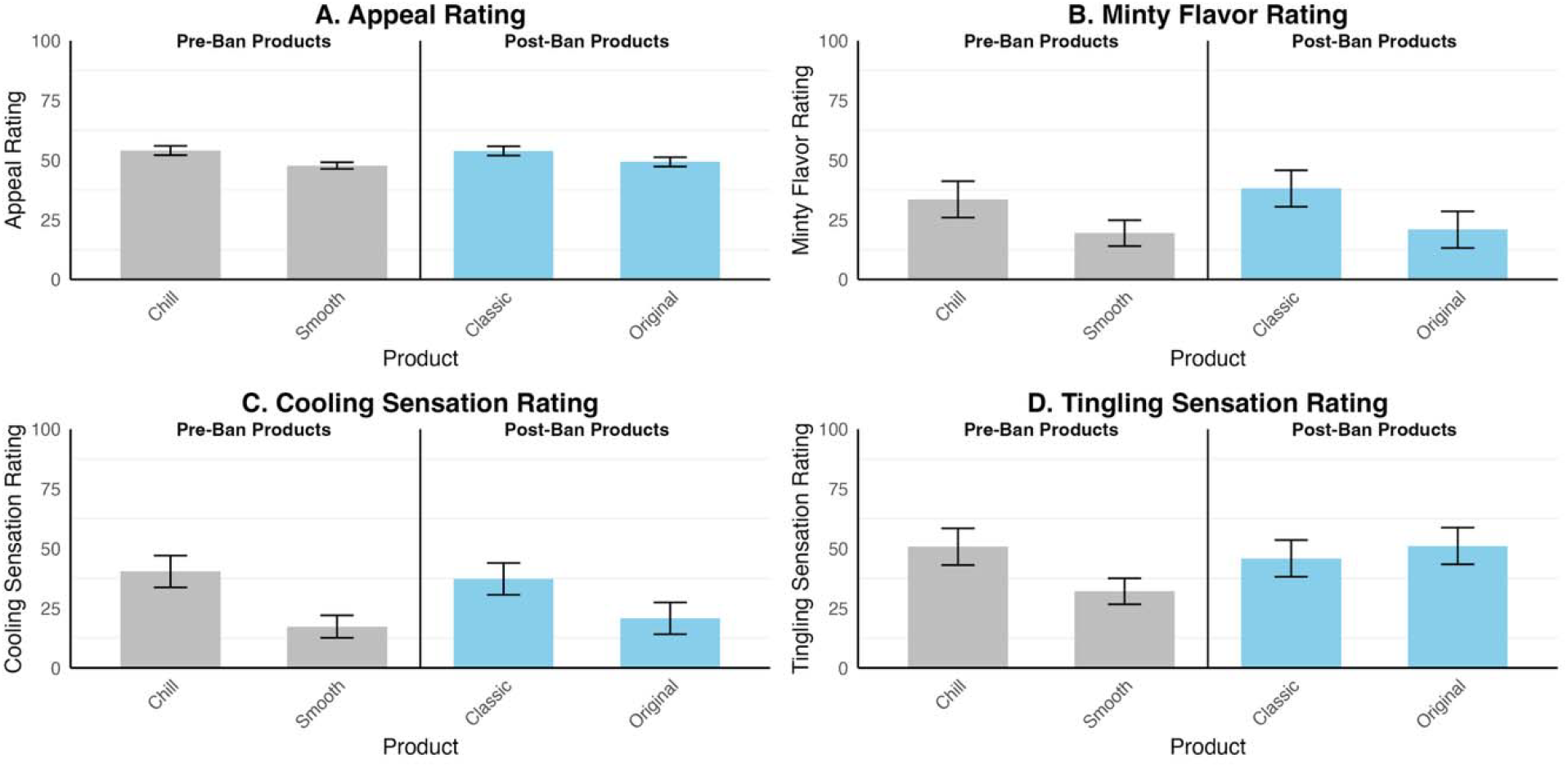
Mean subjective appeal and sensory ratings across ZYN ONPs pre- and post-flavor ban in California *Note*. Panel A shows composite appeal rating, calculated from liking, disliking (reverse-coded), and willingness to use again ratings. Panel B-D displays the ratings for minty flavor, cooling sensation, and tingling sensation. Smooth and Chill represent pre-ban products, while Classic and Original are post-ban products introduced following California’s flavored tobacco ban. Smooth, marketed as a flavorless ONP, was used as the reference product in statistical analyses. Error bars represent ±1 standard error.

## DISCUSSION

Our multidisciplinary study demonstrated that ONP products marketed in California after the flavor ban have similar chemical characteristics, chemosensory receptor activity, and sensory appeal properties among YA’s as pre-ban ONPs. Our results suggest that industry tactics to circumvent flavor ban policies may include overt changes in product branding while maintaining similar appeal and flavor effects, emphasizing the need to address not only the chemical makeup but also branding and package design in tobacco regulations.

Recent regulatory filings further underscore this point. According to the FDA’s 2025 Modified Risk Tobacco Product (MRTP) documentation for ZYN products, it was indicated that ZYN “Smooth” and “Chill” may also be marketed as “Original” and “Classic”, respectively.^31^ Interestingly, this was not noted in FDA’s premarket tobacco product application (PMTA)^32,33^ authorization for marketing 20 ZYN nicotine pouch products. This overlap in product naming highlights how rebranding can complicate post-ban surveillance and enforcement^25^ by obscuring whether changes reflect product reformulation or merely labeling rebranding. Such practices may hinder evaluation of flavor policy effectiveness and allow products to retain their appeal despite legislative restrictions.

Recently, new California legislation revised the definition for “flavored tobacco product” to include products imparting “a cooling sensation”.^17,22^ The continued marketing of ZYN “Classic”, with the addition of synthetic cooling agent WS-3 and its noticeable cooling effect, may violate this law. Prior work^16^ and our current receptor activity findings demonstrate together that, compared to menthol-containing ONPs, WS-3 only containing ONPs (Chill and Classic) displayed higher efficacy for activation of cold/menthol receptor TRPM8, but weak potency and efficacy for the sensory-irritant menthol receptor TRPA1. These receptor activity studies indicate that these WS-3 only containing pre-ban and post-ban ONPs have similar activation profiles, producing robust cooling sensation like menthol-/mint-flavored ONPs, but with reduced sensory irritancy.

Preclinical rodent studies indicate that, like menthol, synthetic cooling agents exhibit analgesic properties and thereby facilitate in reducing the sensory irritation produced by nicotine in ONPs.^34^ In intravenous self-administration models in rats, it was also shown that cooling sensation induced by either cold water, menthol, or synthetic cooling agents can have strong reinforcing effects on nicotine intake.^35^ Together, these findings imply that odorless synthetic cooling agents used in ONPs (e.g., ZYN Chill or Classic) may enhance the perceived sensory experience by reducing irritation and providing a cooling effect, and thereby potentially facilitate initiation and continuation of use.

Clinical and pre-clinical studies show that cooling additives in tobacco products increase appeal and preference among youth,^36,37^ particularly by masking nicotine’s harshness and making use more tolerable. In our study, participants clearly perceived stronger cooling sensations from “Chill” and “Classic” compared with the flavorless “Smooth” and “Original.” This aligns with California’s revised definition of characterizing flavor, which includes “a cooling sensation distinguishable by an ordinary consumer during the consumption of a tobacco product”, suggesting that the cooling reported by participants may meet the criteria of this new definition and therefore could place these products in violation of the ban.^17,22^ In addition, “Classic” was uniquely perceived as more minty despite no detectable menthol, suggesting that WS-3 may contribute to mint/menthol-like effects^16^ that flavor bans are designed to eliminate. While overall appeal ratings did not differ significantly across pre- and post-ban products, this may reflect the high similarity of these formulations.

Indeed, in the e-cigarette literature, synthetic cooling agents added to e-cigarettes have been shown to increase appeal, regardless of product flavor or nicotine concentration,^37^ and increase nicotine vaping and frequency of use among youth.^38^ To our knowledge, these are the first data from a human trial examining appeal of synthetic cooling agents added to ONPs. Our results highlight that synthetic cooling agents may be used to preserve the flavor-like qualities of ostensibly “non-flavored” products, thus, weakening regulations intended to prevent sensory features that make nicotine more tolerable and appealing to youth and young adults.

Presence of synthetic cooling agents in popular ONPs raises toxicity concerns, especially if added at high amounts and/or associated with heavy use, as consumer exposures can exceed safety thresholds set by regulatory agencies.^21^ While FDA has approved several cooling agents, such as WS-3 and WS-23, for use in food, their oral and systemic health effects are unknown. Further, colorful packaging that indicate flavors significantly increase the appeal and use of tobacco products, particularly among young people. Tobacco companies have also intentionally used colors and imagery to manipulate consumer perceptions of products sensory attributes and taste. Tobacco regulations should consider both restricting these additives, since they undermine the public health benefits of flavor bans, and other considerations such as packaging standards (e.g., plain packaging) to reduce the influence of branding elements (e.g., colors, imagery, concept names) that can implicitly signal flavors and increase product appeal despite flavor restrictions.

This study has limitations. First, the human trial may have been underpowered to precisely estimate subjective sensory experiences, yet results were consistent across methods within this rapid, multimethod approach using subjective, chemical and bioassay tools to assess regulatory compliance of emerging products. Second, only “Classic” and “Original” were available for purchase in California and these specific flavors may be limited or unavailable outside that market. Additionally, although Classic was perceived as more minty, we did not test for all possible mint-related compounds (e.g., menthone), and the presence of other unmeasured chemicals may explain this sensory difference. Lastly, though only ZYN brand of products was tested, ZYN is the market-leading ONP brand. Future studies should include other ONP brands to evaluate compliance with flavor restriction policies.

## CONCLUSIONS

Data suggest that the core sensory experience, properties, and chemical characteristics of pre- and post-ban ONPs from a leading brand, remain consistent, and thereby undermine the intended public health gains of flavor-bans regarding reductions in youth appeal, initiation, and health disparities. These findings highlight the enforcement challenges that arise when industry tactics exploit regulatory loopholes through branding strategies and addition of odorless sensory chemicals with attributes similar to characterizing flavors, allowing products to retain their appeal despite legislative restrictions. At a population level, other regulatory agencies could consider following California’s example to close such gaps in regulation which threaten to sustain nicotine uptake among young people and exacerbate inequities in communities already disproportionately affected by tobacco use. This study also demonstrates the importance of multidisciplinary approach of evaluating products to ensure compliance with tobacco regulation.

## Data Availability

All data produced in the present study are available upon reasonable request to the authors.

